# Clinical Profile, Comorbidities, and Outcome of the Unvaccinated and Hospitalized for COVID-19 in Northern Brazil: Retrospective Cohort

**DOI:** 10.1101/2023.06.29.23292037

**Authors:** Ana Lúcia da Silva Ferreira, Daniele Melo Sardinha, Daiane Cristina Viana de Moraes, Maria Raimunda Rodrigues de Oliveira, Mayara Carolina Frazão Viana, Natasha Cristina Oliveira Andrade, Tamires de Nazaré Soares, Ricardo José de Paula Souza e Guimarães, Luana Nepomuceno Gondim Costa Lima, Karla Valéria Batista Lima

## Abstract

Over the course of the pandemic, COVID-19 affected health, the economy and quality of life in Brazil. The worst years for the country were the first and second. There were delays in vaccine purchases for political reasons at the time. The northern region of the country had a higher mortality rate compared to other regions, associated with local vulnerabilities and fragility of surveillance due to geographic and population characteristics. This study aims to investigate the clinical profile, comorbidities, and outcome of unvaccinated people hospitalized for COVID-19 in the state of Pará in 2022. Retrospective cohort epidemiological study, with data from the national epidemiological surveillance of acute and severe respiratory syndromes. Cases reported in 2022 with vaccinated yes or no field and completed doses were included. Only closed cases cure or death were included. We performed a chi-square test on categorical variables and a Mann-Whitney test on numerical variables. We compared vaccinated VS non-vaccinated; we performed the Odds Ratio in the significant variables. We used the SPSS 20.0 software. The study worked with 2,634 cases of COVID-19 hospitalized in the study period, confirmed by RT-PCR (851/32.30%) and (1,784/67.70%) rapid antigen test. The lethality was (778/29.53%), and those vaccinated with two doses were (1,473/55.90%) and those unvaccinated with no dose (1,162/44.10%). Death represents p-<0.001 (HR 1.306 - CI 1.124/1.517) higher risk of the event occurring in the unvaccinated cases, followed by male sex p-0.004 (HR 1.188 - CI 1.058/1.334).. The first cohort in Brazil and in the north of the country to evaluate the clinical profile, comorbidities, and outcome of COVID-19 in hospitalized patients in this Amazon region, which is a region characterized by local vulnerability factors unique to the other regions of Brazil, showed that the unvaccinated were males, younger, with fewer comorbidities, and that they were associated the deaths.

## 1. Introduction

COVID-19 devastated Brazil and the world in the first and second years of the pandemic, especially Brazil, one of the reasons was the delay in purchasing the vaccines, for political reasons at the time, which was denounced by the National Health Council (NHC) as a violation of human rights by the conduct of the Bolsonaro government in the pandemic, which even tried to influence the use of drugs without scientific evidence[1].

With 319,119 confirmed cases, the COVID-19 in Brazil in 2020 peaked in July. The number of deaths also reached 7,453 in July 2020. In 2021, the country faced a wave of greater intensity, with a peak of cases in March, with 533,024 confirmed cases and 21,094 deaths in March 2021 alone. It was the worst phase of the disease because we didn’t have the vaccine. In 2022, we saw the highest number of cases since the pandemic began, but they were mostly mild and only reached 1,283,024 in January 2022. However, the number of deaths was much lower than in previous years, with 6,658 deaths in January 2022, probably due to vaccination[2].

COVID-19 surveillance in Brazil standardized the molecular test for diagnosis in the first year of the pandemic. This was because surveillance for influenza syndromes was already carried out in sentinel units, and all cases of severe acute respiratory syndrome in hospitalized patients and deaths were screened for respiratory viruses. In the second year of the pandemic, diagnosis was strengthened with the introduction of antigen tests, which were essential tools for rapid diagnosis and isolation of patients and for breaking the chain of transmission. However, molecular testing for respiratory viruses has always been mandatory in severe cases[3].

Vaccination started slowly in Brazil in January 2021, initially for the elderly over 90 years old and health professionals, while we faced the second wave with thousands of deaths. Progress was slow and we were only able to vaccinate the entire adult population with the first dose by the end of 2021. Children were vaccinated in early 2022 and infants were not vaccinated, despite the Fizer baby being approved by the National Health Surveillance Agency (ANVISA). The delay in vaccine procurement and slow vaccination by age group may have influenced the mortality of COVID-19 by the end of 2022[4–9].

Studies conducted from 2020 to 2021 on vaccine use and mortality reduction in other countries that started vaccination before Brazil, showed mortality reduction still in 2021, including a reduction in the elderly[10,11]. A systematic review with meta-analysis with 54 studies, showed the Efficacy of the Pooled Vaccine (EPV) against SARS-COV 2 infection was 71% [odds ratio (OR) = 0.29, 95% confidence interval (CI): 0.23-0.36] at the first dose and 87% (OR = 0.13, 95% CI: 0.08-0.21) at the second dose. The EPV for preventing hospitalization due to COVID-19 infection was 73% (OR = 0.27, 95% CI: 0.18-0.41) at the first dose and 89% (OR = 0.11, 95% CI: 0.07-0.17) at the second dose. Regarding the type of vaccine, mRNA-1273 and combined studies at the first dose and ChAdOx1 and mRNA-1273 at the second dose showed higher efficacy in preventing infection. Regarding COVID-19-related mortality, EPV was 68% (HR = 0.32, 95% CI: 0.23-0.45) at the first dose and 92% (HR = 0.08, 95% CI: 0.02-0.29) at the second dose[12]. Vaccines are the best strategies to control the pandemic and reduce mortality, especially among the elderly [13].

The northern region of the country had a higher mortality rate compared to other regions, associated with local vulnerabilities and fragility of surveillance due to geographic and population characteristics. One study described the predictors of deaths in the state of Pará in the first year of the pandemic, and showed that the predictors of lethality were invasive ventilation (OR 6,627; CI 5,780-7,597), other pneumopathy (OR 1.901; CI 1.439-2.510), dyspnea (OR 1.899; CI 1.737-2.076), immunodeficiency (OR 1. 905; CI 1.493-2.431), hospitalized in ICU (OR 1.764; CI 1.588-1.959), chronic kidney disease (OR 1.753; CI 1.396-2.203), diabetes mellitus (OR 1.210; CI 1.108-1.321), and male gender (OR 1.198; CI 1.111-1.293). And it showed that those vaccinated against influenza were associated with survivors[14].

No study has evaluated the profile of those hospitalized for COVID-19 in the third year of the pandemic in this region of Brazil, because it is worth noting that denialism greatly influenced non-adherence to COVID-19 vaccines. Assessing the factors associated with non-vaccination in hospitalized cases will guide public health to more effective strategies for COVID-19 vaccine campaigns and improve the immunization scenario and reduce severe cases and deaths in this region. Thus, we question: What are the clinical profile, comorbidities, and outcomes of unvaccinated people hospitalized for COVID-19 in the state of Pará in 2022?

## 2. Materials and Methods

### 2.1. Type of study

The present study is a retrospective cohort with regional data from the surveillance of acute and severe respiratory syndromes in the state of Pará, Brazil. It was carried out in the year 2022. According to the guidelines of the Strengthening the reporting of observational studies in epidemiology (STROBE) [15].

### 2.2. Study location

The study was conducted in the state of Pará, which is located in the northern region of Brazil (figure 1). Pará is the second largest state in the country in territorial extension, with an area of 1,245,870.798 km^2^, has an estimated population for 2020 of 8,690,745 inhabitants, and presents a Human Development Index (HDI) of 0.646. The state has six Mesoregions comprised of 22 Microregions, in a total of 144 municipalities, and has the city of Belém as its capital. The territory of Pará is composed of the largest tropical forest in the world, the Amazon. The relief is low and flat; 58% of the territory is below 200 meters. Altitudes above 500 meters are found in the following mountain ranges: Serra dos Carajás, Serra do Cachimbo, and Serra do Acari. [16].

**Figure 1.**
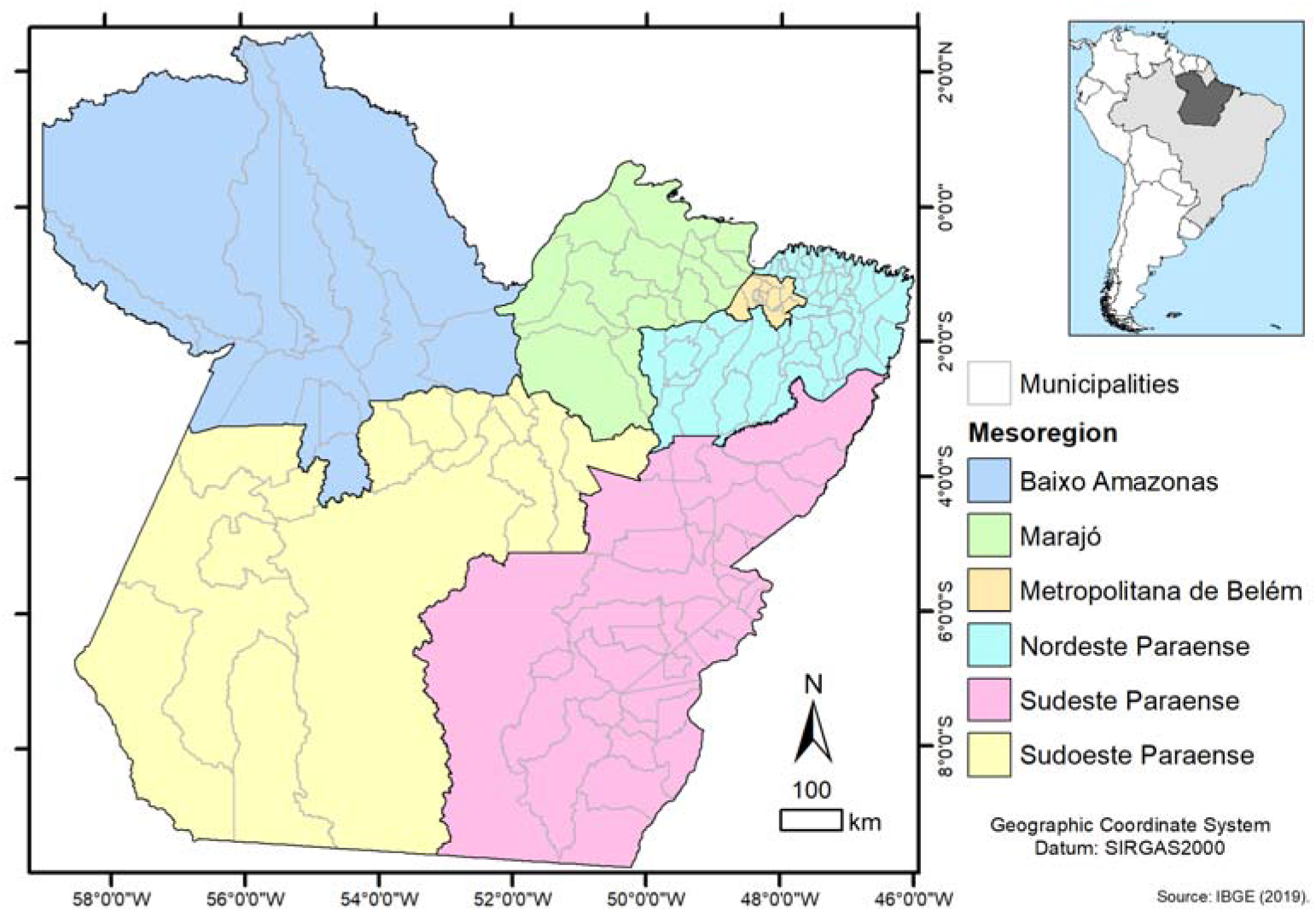
Geographic location of the state of Pará and its mesoregions. Source: Sardinha, [17].

**Figure 2.**
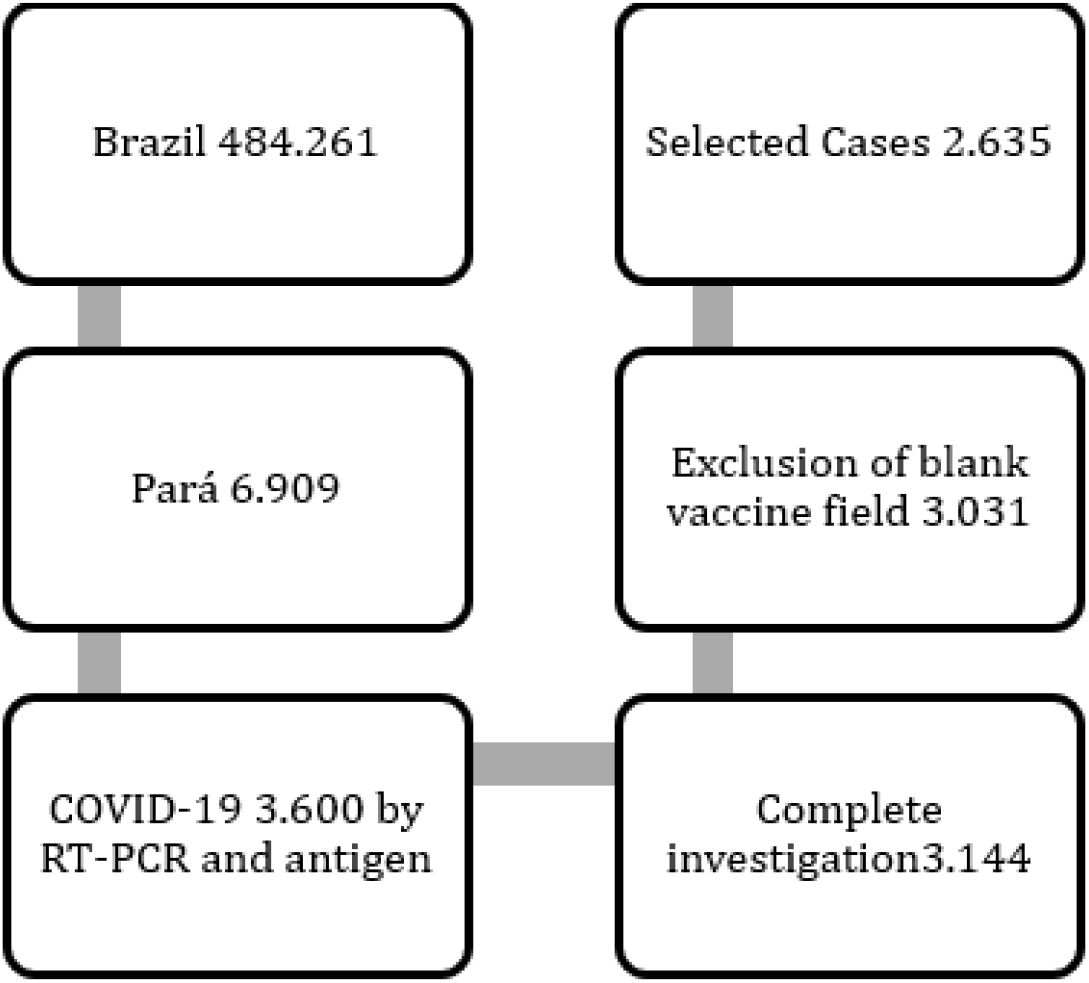
Selection of participants. Source: Authors’ research.

### 2.2. Study population and source

The study population was all cases reported and confirmed by RT-PCR and rapid antigen testing for COVID-19 in the Influenza Epidemiological Surveillance Information System platform SIVEP-GRIPE (https://sivepgripe.saude.gov.br/sivepgripe/login.html?1). In the period January 01, 2022, to November 09, 2022. The following flow details the selection of participants.

### 2.3. Inclusion Criteria

All confirmed cases, with the vaccination field of COVID-19 filled in, the cases that had yes in the vaccine field should have the dates of the first and second dose, final classification, and evolution filled in.

### 2.4. Exclusion Criteria

Cases with only one vaccine dose completed and no vaccine date.

### 2.5. Case Definition

Severe Acute Respiratory Syndrome (SARS) is defined as individuals with Influenza Syndrome (RSS) who have: dyspnea/respiratory distress or persistent chest pressure or O2 saturation less than 95% on room air or bluish coloration of the lips or face. (SG: Individual with an acute respiratory condition characterized by at least two (2) of the following signs and symptoms: fever (even if referred), chills, sore throat, headache, cough, runny nose, smell or taste disturbances). For notification in SIVEP-GRIPE, should be considered the cases of SARS hospitalized or deaths by SARS independent of hospitalization[18].

The dependent variable of this cohort was cases of unvaccinated (1,162 exposed) and vaccinated (1,473 unexposed) SARS due to COVID-19. A comparison of the clinical, comorbidity, and outcome variables of COVID-19 SARS in the exposed (unvaccinated) and unexposed (vaccinated) groups was performed.

### 2.6. Data Analysis

The database was made available in Excel 2019 format with the variables corresponding to the SIVEP-GRIPE notification form, which is composed of 80 variables, referring to sociodemographic and clinical-epidemiological data. The variables worked and extracted according to the form were: gender (item 8), age (item 10), signs and symptoms (item 35), risk factors/comorbidities (item 36), admitted to ICU (item 47), used ventilatory support (item 50) and evolution (item 74). SIVEP-GRIPE notification form[18].

For the statistics, the absolute and relative frequencies of all cases were calculated, and to investigate the predictor variables in unvaccinated cases, the comparison in a univariate model was performed in the categorical variables by 2×2 table and the Chi-square test between vaccinated and unvaccinated cases, in values less than <5 the Fisher’s exact test was performed, and in significant cases, the Odds Ratio (OR) was also performed.

For the numerical variable age, a normality test was performed, the Kolmogorov-Smirnov test, which was significant (<0.001), thus the Mann-Whitney test was applied to age, to compare the median of the unvaccinated and vaccinated.

We performed COX regression, univariate and multivariate a final model with only the variables that were significant. Cox regression builds a predictive model for time-to-event data. The model produces a survival function that predicts the probability that the event of interest has occurred at a given time t for given values of the predictor variables. The form of the survival function and the regression coefficients for the predictors are estimated from observed subjects; the model can then be applied to new cases that have measures for the predictor variables. Note that information from censored subjects, that is, those who do not experience the event of interest during the observation time, makes a useful contribution to model estimation. The time variable was the day’s counting from the date of notification until the outcome, cure or death. We performed with the dependent variable not vaccinated against COVID-19, and set up with the remaining independent variables, referring to the epidemiological, clinical profile, comorbidities, and disease outcome.For all statistical analyses we used the Statistical Package for the Social Sciences (SPSS) version 26.0. The alpha level of significance for all analyses was <0.05.

### 2.7. Ethical Aspects

And to meet the ethical aspects from Resolution No. 466 of 12 December 2012 that directs the study in the principles of bioethics, emphasizing respect for human dignity, freedom, and autonomy of the human being, also integrating non-maleficence, beneficence, justice, and equity among others, to ensure rights and duties to all involved in research. The project was approved on 11/16/2020 by the Ethics and Research Committee of the Marco School Health Center of the Universidade do Estado do Pará - UEPA. Consubstantiated Opinion No. 4.399.970. Which is part of this multicenter project “SOCIAL, EPIDEMIOLOGICAL AND SPACIAL ANALYSIS OF COVID-19 IN THE STATE OF PARÁ DURING THE PANDEMIA” of the postgraduate program in Parasite Biology in the Amazon of the University of Pará State and the Evandro Chagas Institute (UEPA-IEC).

## 3. Results

The study worked with 2,634 cases of COVID-19 hospitalized in the study period, confirmed by RT-PCR (851/32.30%) and (1,784/67.70%) rapid antigen test. The lethality was (778/29.53%), and those vaccinated with two doses were (1,473/55.90%) and those unvaccinated with no dose (1,162/44.10%).

We present the first and second doses of the vaccine by production laboratory, which were used in Brazil. AstraZeneca was the most frequent in hospitalized patients, in the 1st dose with 52.61% and 2nd with 52.31 (table1). About the outcomes of cure and death, CoronaVac presented the highest lethality with 33.80%(table 2). About Janssen and those hospitalized were lower in number, which may be associated with the start of vaccination with Janssen only in June 2021 and prioritizing the most distant regions of the state.

**Table 1.** First and second doses of the COVID-19 vaccine by production laboratory, of vaccinees hospitalized in the state of Pará in 2022.

| Vaccine Laboratory | 1st dose % | 2st dose % |
| --- | --- | --- |
| AstraZeneca | 52.61 | 52.31 |
| CoronaVac | 38.82 | 38.40 |
| Janssen | 0.24 | 0.12 |
| Pfizer/Biontech | 8.32 | 9.17 |
| **Total** | **100.00** | **100.00** |

Source: SIVEP-GRIPE.

**Table 2.** First and second doses of the COVID-19 vaccine by laboratory and outcome, of vaccinees hospitalized in the state of Pará in 2022.

| 1st dose | Survivor % | Death % | Total % |
| --- | --- | --- | --- |
| AstraZeneca | 71.25 | 28.75 | 52.61 |
| CoronaVac | 66.20 | 33.80 | 38.82 |
| Janssen | 100.00 | 0.00 | 0.24 |
| Pfizer/Biontech | 81.02 | 18.98 | 8.32 |
| **Total** | **70.17** | **29.83** | **100.00** |
| 2nd dose | Survivor % | Death % | Total % |
| AstraZeneca | 71.43 | 28.57 | 52.31 |
| CoronaVac | 66.77 | 33.23 | 38.40 |
| Janssen | 100.00 | 0.00 | 0.12 |
| Pfizer/Biontech | 76.82 | 23.18 | 9.17 |
| **Total** | **70.17** | **29.83** | **100.00** |

Source: SIVEP-GRIPE.

The median age overall was 56 years, of those vaccinated 65 and of those not vaccinated 45, a lower median age was associated with the unvaccinated cases (p-<0.001). About the sex, the male was associated with the unvaccinated (p-0.011- OR 1.202- 95% CI 1.030/1.402).. Fever was associated with the unvaccinated (p-0.013- OR 1.202- 95% CI 1.025/1.408), and vomiting (p-0.021- OR 1.277- 95% CI- 1.016/1.607). Other signs and symptoms had an association with that vaccinated, dyspnea (p-0.001- OR 0.785- 95% CI 0.671/0.918) and O2 saturation <95% (p-0.007- OR 0.819- 95% CI 0.701/0.957). When comparing comorbidities between vaccinated and unvaccinated, significance was associated with vaccinated, showing that individuals with comorbidities are vaccinated for COVID-19, such as in chronic cardiovascular disease (p-<0. 001- OR 0.476- 95% CI 0.396/0.573), chronic liver disease (p-0.049- OR 0.315- 95% CI 0.089/1.119), asthma (p-0.014- OR 0.418- 95% CI 0.196/0.892), diabetes mellitus (p<0. 001- OR 0.515- 95% CI 0.418/0.635), chronic neurological disease (p-0.014- OR 0.579- 95% CI 0.361/0.928), chronic kidney disease (p-0.001- OR 0.480- 95% CI 0.303/0.760). At outcome, ICU admission (p-0.009-OR 1.494-CI 1.105-2.019) and death (p-0.046-OR 1.442-CI 1.163-1.788) were associated with those not vaccinated against COVID-19.(table 3).

**Table 3.**
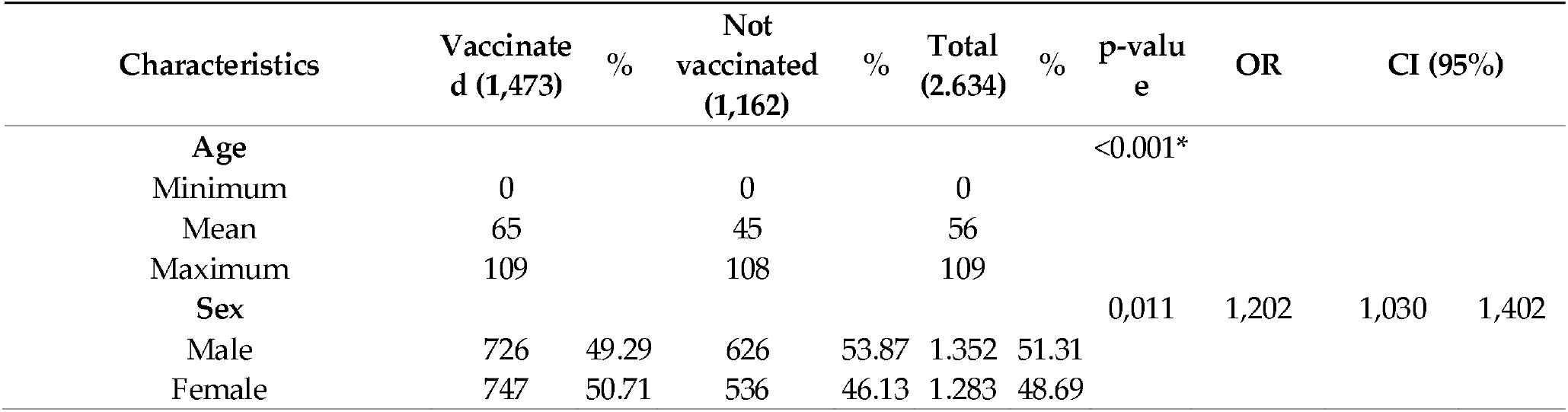
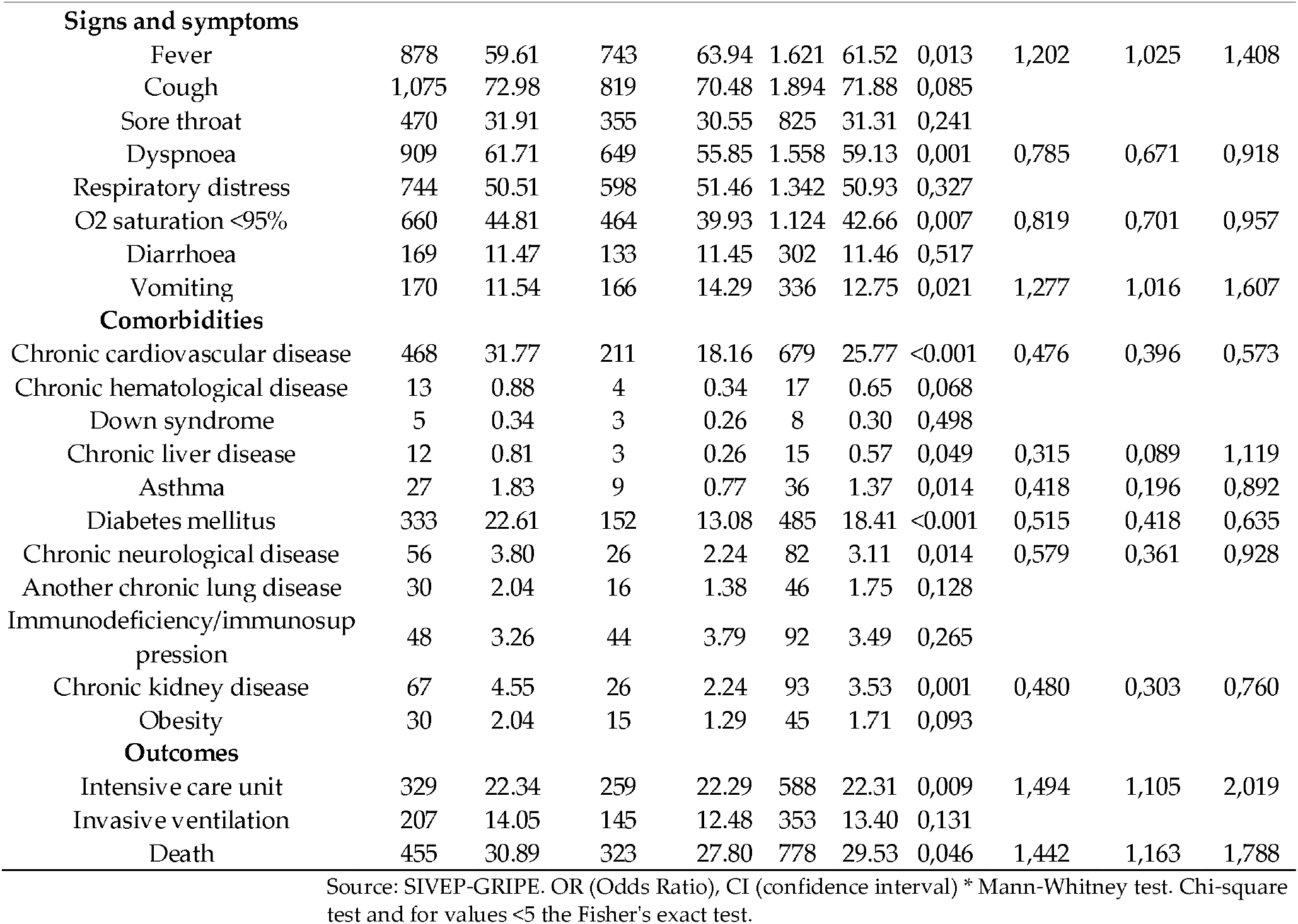
Epidemiological, clinical characteristics, comorbidities, and outcome associated with unvaccinated people hospitalized for COVID-19 in Pará State in 2022.

| Characteristics | Vaccinate<br>d (1,473) | % | Not<br>vaccinated<br>(1,162) | % | Total<br>(2.634) | % | p-value | OR | CI (95%) |
| --- | --- | --- | --- | --- | --- | --- | --- | --- | --- |
| **Age** |  |  |  |  |  |  | <0.001* |  |  |
| Minimum | 0 |  | 0 |  | 0 |  |  |  |  |
| Mean | 65 |  | 45 |  | 56 |  |  |  |  |
| Maximum | 109 |  | 108 |  | 109 |  |  |  |  |
| **Sex** |  |  |  |  |  |  | 0,011 | 1,202 | 1,030 1,402 |
| Male | 726 | 49.29 | 626 | 53.87 | 1.352 | 51.31 |  |  |  |
| Female | 747 | 50.71 | 536 | 46.13 | 1.283 | 48.69 |  |  |  |

|  |  |  |  |  |  |  |  |  |  |  |
| --- | --- | --- | --- | --- | --- | --- | --- | --- | --- | --- |
| **Signs and symptoms** |  |  |  |  |  |  |  |  |  |  |
| Fever | 878 | 59.61 | 743 | 63.94 | 1.621 | 61.52 | 0,013 | 1,202 | 1,025 | 1,408 |
| Cough | 1,075 | 72.98 | 819 | 70.48 | 1.894 | 71.88 | 0,085 |  |  |  |
| Sore throat | 470 | 31.91 | 355 | 30.55 | 825 | 31.31 | 0,241 |  |  |  |
| Dyspnoea | 909 | 61.71 | 649 | 55.85 | 1.558 | 59.13 | 0,001 | 0,785 | 0,671 | 0,918 |
| Respiratory distress | 744 | 50.51 | 598 | 51.46 | 1.342 | 50.93 | 0,327 |  |  |  |
| O2 saturation <95% | 660 | 44.81 | 464 | 39.93 | 1.124 | 42.66 | 0,007 | 0,819 | 0,701 | 0,957 |
| Diarrhoea | 169 | 11.47 | 133 | 11.45 | 302 | 11.46 | 0,517 |  |  |  |
| Vomiting | 170 | 11.54 | 166 | 14.29 | 336 | 12.75 | 0,021 | 1,277 | 1,016 | 1,607 |
| **Comorbidities** |  |  |  |  |  |  |  |  |  |  |
| Chronic cardiovascular disease | 468 | 31.77 | 211 | 18.16 | 679 | 25.77 | <0.001 | 0,476 | 0,396 | 0,573 |
| Chronic hematological disease | 13 | 0.88 | 4 | 0.34 | 17 | 0.65 | 0,068 |  |  |  |
| Down syndrome | 5 | 0.34 | 3 | 0.26 | 8 | 0.30 | 0,498 |  |  |  |
| Chronic liver disease | 12 | 0.81 | 3 | 0.26 | 15 | 0.57 | 0,049 | 0,315 | 0,089 | 1,119 |
| Asthma | 27 | 1.83 | 9 | 0.77 | 36 | 1.37 | 0,014 | 0,418 | 0,196 | 0,892 |
| Diabetes mellitus | 333 | 22.61 | 152 | 13.08 | 485 | 18.41 | <0.001 | 0,515 | 0,418 | 0,635 |
| Chronic neurological disease | 56 | 3.80 | 26 | 2.24 | 82 | 3.11 | 0,014 | 0,579 | 0,361 | 0,928 |
| Another chronic lung disease | 30 | 2.04 | 16 | 1.38 | 46 | 1.75 | 0,128 |  |  |  |
| Immunodeficiency/immunosuppression | 48 | 3.26 | 44 | 3.79 | 92 | 3.49 | 0,265 |  |  |  |
| Chronic kidney disease | 67 | 4.55 | 26 | 2.24 | 93 | 3.53 | 0,001 | 0,480 | 0,303 | 0,760 |
| Obesity | 30 | 2.04 | 15 | 1.29 | 45 | 1.71 | 0,093 |  |  |  |
| **Outcomes** |  |  |  |  |  |  |  |  |  |  |
| Intensive care unit | 329 | 22.34 | 259 | 22.29 | 588 | 22.31 | 0,009 | 1,494 | 1,105 | 2,019 |
| Invasive ventilation | 207 | 14.05 | 145 | 12.48 | 353 | 13.40 | 0,131 |  |  |  |
| Death | 455 | 30.89 | 323 | 27.80 | 778 | 29.53 | 0,046 | 1,442 | 1,163 | 1,788 |

Source: SIVEP-GRIPE. OR (Odds Ratio), CI (confidence interval) \* Mann-Whitney test. Chi-square test and for values <5 the Fisher's exact test.

In the final multivariate survival model, by COX regression, we identified the hazard ratio in the variables that form significant for the dependent variable unvaccinated against COVID-19. Death represents p-<0.001 (HR 1.306 - CI 1.124/1.517) higher risk of the event occurring in the unvaccinated cases, followed by male sex p-0.004 (HR 1.188 - CI 1.058/1.334). The other significant variables in the multivariate model were negatively associated, i.e., those vaccinated were associated with older age, having comorbidities and fever, cough and invasive ventilation, showing that those vaccinated have older age, more comorbidities, fever and cough symptoms, and higher rates of invasive ventilation. The non-vaccinated are young, male and have fewer comorbidities, and a higher risk of death (Table 4).

**Table 4.** Final multivariate survival model, COX regression, hazard ratio for unvaccinated COVID-19 in hospitalized patients in the northern region of Brazil in 2022.

| Variables | Univariate |  | 95,0% CI |  | Multivariate |  | 95,0% CI |  |
| --- | --- | --- | --- | --- | --- | --- | --- | --- |
|  | p-value | HR adjusted | Lower | Upper | p-value | HR adjusted | Lower | Upper |
| Death | <0,001 | 1,317 | 1,128 | 1,538 | <0,001 | 1,306 | 1,124 | 1,517 |
| UCI | 0,650 | 0,954 | 0,777 | 1,170 |  |  |  |  |
| Invasive ventilation | 0,111 | 0,815 | 0,634 | 1,048 | 0,008 | 0,773 | 0,639 | 0,936 |
| Fever | 0,097 | 0,892 | 0,779 | 1,021 | 0,033 | 0,867 | 0,761 | 0,988 |
| Cough | 0,102 | 0,890 | 0,774 | 1,023 | 0,030 | 0,861 | 0,753 | 0,985 |
| Sorethroat | 0,267 | 0,925 | 0,805 | 1,062 |  |  |  |  |
| Dyspnoea | 0,302 | 0,931 | 0,813 | 1,066 |  |  |  |  |
| Respiratory distress | 0,291 | 0,931 | 0,816 | 1,063 |  |  |  |  |
| O2 saturation 95% | 0,746 | 1,024 | 0,888 | 1,180 |  |  |  |  |
| Diarrhoea | 0,665 | 0,958 | 0,789 | 1,163 |  |  |  |  |
| Vomiting | 0,745 | 0,971 | 0,814 | 1,159 |  |  |  |  |
| Male | 0,006 | 1,180 | 1,049 | 1,326 | 0,004 | 1,188 | 1,058 | 1,334 |
| Age | <0,001 | 0,985 | 0,983 | 0,987 | <0,001 | 0,985 | 0,982 | 0,987 |
| Chronic cardiovascular disease | 0,271 | 0,910 | 0,769 | 1,077 | 0,016 | 0,892 | 0,759 | 1,048 |
| Chronic hematological disease | 0,266 | 0,571 | 0,213 | 1,534 |  |  |  |  |
| Down syndrome | 0,496 | 1,486 | 0,475 | 4,647 |  |  |  |  |
| Chronic liver disease | 0,559 | 0,712 | 0,228 | 2,225 |  |  |  |  |
| Asthma | 0,348 | 0,728 | 0,375 | 1,413 |  |  |  |  |
| Diabetes mellitus | 0,376 | 0,919 | 0,763 | 1,108 |  |  |  |  |
| Chronic neurological disease | 0,773 | 0,944 | 0,639 | 1,396 |  |  |  |  |
| Another chronic lung disease | 0,651 | 0,890 | 0,538 | 1,472 |  |  |  |  |
| Immunodeficiency/immunosuppression | 0,056 | 0,739 | 0,542 | 1,007 | 0,035 | 0,719 | 0,528 | 0,978 |
| Chronic kidney disease | 0,011 | 0,599 | 0,404 | 0,888 | 0,008 | 0,587 | 0,396 | 0,868 |
| Obesity | 0,529 | 0,847 | 0,506 | 1,420 |  |  |  |  |

Source: SIVEP-GRIPE. HR (Hazard Ratio), CI (Confidence Interval).

## 4. Discussion

This study analyzed the epidemiological profile factors, comorbidities, and the outcome of COVID-19 in vaccinated and unvaccinated hospitalized patients by COVID-19. To our knowledge, it is the first cohort in the northern region of Brazil with the dependent variable unvaccinated. We show that overall lethality was 29.53%, lower than a study that analyzed lethality in the first year of the pandemic before vaccination in the state of Pará, which was 42.47%. [14]. Evidence that the vaccine reduced the lethality rate in the 2022 period when compared to 2020.

A study in the United States estimated the risk of hospitalizations among vaccinated and unvaccinated patients from a national database and showed that the rates of hospitalizations in unvaccinated cases for COVID-19 among the infected were 10 times higher than among vaccinated patients, and the study underscored the importance of full-dose and booster vaccination for reducing hospitalization and deaths. [19]. In our cohort, we did not measure the risk of hospitalizations, because we only analyzed those who were hospitalized. Other research has associated the lower risk of mortality from non-COVID-19 causes in those vaccinated for COVID-19 and has emphasized the safety of COVID-19 vaccines [20].

An analysis of vaccinated and unvaccinated healthcare professionals specifically infected with the gamma variant in early 2021 in a tertiary care hospital in São Paulo, Brazil, showed that the most common symptoms in vaccinated patients on multivariate after adjustment for multiple comparisons, hyposmia/anosmia (OR = 0.304, adj p < 0.001) and dysgeusia (OR = 0.385, adj p = 0.011) were the only symptoms significantly associated with the gamma variant [21]. However, in our study, we analyzed hospitalized severe cases in the year 2022, when the variant of SARS-CoV-2 in the reported cases was not known in the database.

A cohort in Italy compared COVID-19 waves before and after the vaccine and found that the vaccinated group was older and had comorbidities [22]. Another cohort at Aga Khan University Hospital (AKUH) of 434 patients. For unvaccinated patients, there was a significant increase in the number of intensive care admissions (p-value 0.043). The unvaccinated patients had significantly higher serum levels of procalcitonin, ferritin, LDH, and D-dimer. About 5.3% (n = 23) of patients required invasive ventilation, which was more common in unvaccinated patients (p-value 0.04). Overall, the mortality rate was 12.2% (n = 53) and was higher (16.2%, n = 39) in unvaccinated patients compared with fully vaccinated patients (6.1%, n = 10, p-value 0.006)[23]. Another survey of hospitalized patients showed that unvaccinated patients required mechanical ventilation more frequently (8.5%) than vaccinated patients, in whom the likelihood of mechanical ventilation decreased with increasing doses (8.7%, 2.8%, 0%)[24]. This is similar to our results on the association of deaths in unvaccinated cases.

A survey that evaluated in-hospital mortality of COVID-19 patients by vaccination status using a retrospective cohort study, in Northern California. Of the 7,305 patients, 1,463 (20.0%) were complete, 138 (1.9%) were partial, and 5,704 (78.1%) were unvaccinated. The fully vaccinated were older than the partial or unvaccinated (71.0, 63.0, and 54.0 years, respectively, p < 0.001) with more comorbidities (Comorbidity Scores 33.0, 22.0, and 10.0, p < 0.001) and immunosuppressants (11.5%, 8.7%, and 3.0%, p < 0.001) or chemotherapy exposure (2.8%, 0.7%, and 0.4%, p < 0.001). Fewer fully vaccinated patients died compared to the corresponding unvaccinated patients (9.0% vs. 16.3%, p < 0.0001). Fully vaccinated patients were less likely to die compared with unvaccinated patients [25]. Another survey identified that those vaccinated with the full regimen were older with comorbidities [26]. Similar to our results, however, we showed higher lethality.

Research has shown vaccine benefits also in fully vaccinated pregnant women infected with SARS-CoV-2 during the Omicron wave had a milder disease and were less likely to require oxygen supplementation and intensive care compared with their unvaccinated counterparts [27].

Our results associated older age with those vaccinated against COVID-19, but when adjusting for age and sex, deaths were associated with those unvaccinated, highlighted in a survey in Spain that analyzed vaccinated and unvaccinated elderly >85 who were hospitalized for COVID-19 and showed a reduction in mortality in this age group in vaccinated cases. This decline may be explained by the greater availability of hospital resources and more effective treatments as the pandemic progressed, although other factors, such as changes in the virulence of SARS-CoV-2, cannot be ruled out [28]. A study analyzed hospitalizations for COVID-19 in the Delta variant period (Jan to Aug 2021) in the United States and concluded that lower vaccination coverage in adults aged 18 to 49 likely contributed to the increase in hospitalized patients during the Delta period. Vaccination against COVID-19 is critical for all eligible adults, including adults under age 50 who have relatively low vaccination rates compared to older adults[29].

We did not delve into the analysis by vaccine type due to the high risk of bias because it is surveillance data, however Pfizer/Biotench in the two doses had the lowest lethality in the hospitalized cohort, we did not include booster doses. One study evaluated Pfizer/Biotench vaccine booster doses and found that at least 5 months after a second dose had 90% lower mortality due to COVID-19 than participants who did not receive a booster [30]. The low numbers vaccinated with Janssen may be associated with the vaccine only starting in June 2021 in the state of Pará, which started with the municipalities farthest from the metropolitan region[31].

One study has already shown that seropositivity for SARS-CoV-2 in unvaccinated individuals has an impact on the reduction of severity and death from COVID-19 infection[32]. Another study found no severe symptoms in patients who were previously seropositive for SARS-CoV-2 in COVID-19[33]. This could be a protective factor in patients who are not vaccinated. However, in our study, the epidemiological surveillance database does not contain information on pre-existing seropositivity in unvaccinated cases.

Vaccination is the main strategy to fight infectious diseases, and it is no different in COVID-19; several studies have shown a reduction in hospitalization and death from the disease [34–36]. Thus, promoting vaccination is the best choice for countries to reduce the impacts of COVID-19.

This study warns that despite being younger and with fewer comorbidities, COVID-19 has worse outcomes in these cases, increasing the risk of death. Thus, vaccination is the best strategy to reduce hospitalizations and deaths from COVID-19, and effective strategies for vaccine adherence must be developed because the current challenge is the fake news and negationist, political, and even religious positioning[37–39].

We highlight the limitation of the study and the risk of bias because we are dealing with secondary data from the national SARS surveillance database; however, the data in 2022 are much better regarding the quality of the information when compared to the first year of the pandemic, since there were fewer cases and the teams were already trained in the notification and investigation of SARS in the surveillance system. Another limitation was the lack of information on the dominant variants because the laboratory analysis of surveillance in Brazil is limited only to molecular testing, without information on genotyping. The high number of multiple vaccinations, which makes it difficult to interpret the overall benefit, is another limitation of the study.

We also highlight as a limitation of a retrospective cohort study with secondary data from epidemiological surveillance, firstly there may be divergence in data collection by region and state of the country, because the quality of data depends directly on local surveillance and health institutions, our region is already considered vulnerable and difficult area to perform surveillance by geographical characteristics. Another point is the socioeconomic differences between the states of the country, since states with better financial conditions have better access to health care services with higher quality, which can influence the outcome healing or death, and vaccination. Third point is the results do not apply to long-term protection against COVID-19 in vaccinated patients, other studies should be conducted in the long term to verify the benefits of the vaccine and even the need for doses in campaigns per year.

Discussing the profile of the unvaccinated allows alerting vaccine campaigns to be directed to this population with greater effectiveness regarding acceptance since the state of Pará continues vaccinating in all health posts in the region. The study is also a warning because it associates deaths with the unvaccinated, although the clinical management in 2022 is already better directed to the severe cases of COVID-19. Vaccination is our best strategy to fight the pandemic by COVID-19.

## 5. Conclusions

The first cohort in Brazil and in the north of the country to evaluate the clinical profile, comorbidities, and outcome of COVID-19 in hospitalized patients in this Amazon region, which is a region characterized by local vulnerability factors unique to the other regions of Brazil, showed that the unvaccinated were men, younger, with fewer comorbidities, and that they were associated the deaths.

We show that hospitalized people unvaccinated for COVID-19, die more from COVID-19 than people vaccinated with two doses, despite advances in clinical management of the disease in 2022, which reinforces the importance of COVID-19 vaccines as an international public health strategy for pandemic control.

## Data Availability

Request the data for the corresponding author.

## Author Contributions

Conceptualization, A.L. da S.F. and D.M.S.; methodology, D.M.S.; software, D.M.D. and T de NS; validation, R.J. de S. e G.; formal analysis, D.M.S..; investigation and resources, A.L. da S.F., D.C.V de M., M.R.R de O., M.C.F.V., and N.C.O.A.; data curation, D.M.S.; writing—original draft preparation, D.M.S; writing—review and editing, A.L. da S.F.; visualization, supervision and project administration, L.N.G.C.L. and K.V.B.L. All authors have read and agreed to the published version of the manuscript.

## Funding

Conselho Nacional de Desenvolvimento Científico e Tecnológico (CNPQ), Coordenação de Aperfeiçoamento de Pessoal de Nível Superior (CAPES), Instituto Evandro Chagas (IEC) and Fundação Amazônia de Amparo an Estudos e Pesquisas (FAPESPA).

## Institutional Review Board Statement

To meet the ethical aspects from Resolution No. 466 of 12 December 2012 that directs the study in the principles of bioethics, emphasizing respect for human dignity, freedom, and autonomy of the human being, also integrating non-maleficence, beneficence, justice, equity among others, to ensure rights and duties to all involved in research. The project was approved on 11/16/2020 by the Ethics and Research Committee of the Marco School Health Center of the Universidade do Estado do Pará - UEPA. Consubstantiated Opinion No. 4.399.970. Which is part of this multicenter project “SOCIAL, EPIDEMIOLOGICAL AND SPACIAL ANALYSIS OF COVID-19 IN THE STATE OF PARÁ DURING THE PANDEMIA” of the post-graduate program in Parasite Biology in the Amazon of the University of Pará State and the Evandro Chagas Institute (UEPA-IEC).

## Informed Consent Statement

Informed Consent Statement: Patient consent was waived due to REASON (retrospective data from regional epidemiological surveillance, whereby personal patient information such as name, phone number, and address was excluded).

## Data Availability Statement

Only by e-mail request from the corresponding author.

## Acknowledgments

Instituto Evandro Chagas.

## Conflicts of Interest

The authors declare no conflict of interest.

## Notes

### Competing Interest Statement

The authors have declared no competing interest.

### Funding Statement

Conselho Nacional de Desenvolvimento Cientifico e Tecnologico (CNPQ), Coordenacao de Aperfeicoamento de Pessoal de Nivel Superior (CAPES), Instituto Evandro Chagas (IEC) and Fundacao Amazonia de Amparo an Estudos e Pesquisas (FAPESPA).

### Author Declarations

The project was approved on 11/16/2020 by the Ethics and Research Committee of the Marco School Health Center of the Universidade do Estado do Pará - UEPA. Consubstantiated Opinion No. 4.399.970.

